# Willingness to seek laboratory testing for SARS-CoV-2 with home, drive-through, and clinic-based specimen collection locations

**DOI:** 10.1101/2020.05.06.20093005

**Authors:** Aaron J. Siegler, Eric Hall, Nicole Luisi, Maria Zlotorzynska, Gretchen Wilde, Travis Sanchez, Heather Bradley, Patrick S. Sullivan

## Abstract

**Background:** SARS-CoV-2 virus testing for persons with COVID-19 symptoms, and contact tracing for those testing positive, will be critical to successful epidemic control. Willingness of persons experiencing symptoms to seek testing may determine the success of this strategy.

**Methods:** A cross-sectional, online survey in the United States measured willingness to seek testing if feeling ill under different specimen collection scenarios: home-based saliva, home-based swab, drive-through facility swab, and clinic-based swab. Instructions clarified that home-collected specimens would be mailed to a laboratory for testing. We presented similar willingness questions regarding testing during follow-up care.

**Results:** Of 1435 participants, comprising a broad range of sociodemographic groups, 92% were willing to test with a home saliva specimen, 88% with home swab, 71% with drive-through swab, and 60% with clinic collected swab. Moreover, 68% indicated they would be more likely to get tested if there was a home testing option. There were no significant differences in willingness items across sociodemographic variables or for those currently experiencing COVID-19 symptoms. Results were nearly identical for willingness to receive testing for follow-up COVID-19 care.

**Conclusions:** We observed a hierarchy of willingness to test for SARS-CoV-2, ordered by the degree of contact required. Home specimen collection options could result in up to one-third more symptomatic persons seeking testing, facilitating contact tracing and optimal clinical care. Remote specimen collection options may ease supply chain challenges and decrease the likelihood of nosocomial transmission. As home specimen collection options receive regulatory approval, they should be scaled rapidly by health systems.

## INTRODUCTION

A central component of COVID-19 disease containment strategies will be scaled-up testing and self-isolation/quarantine as applicable.^1,2^ This strategy requires active identification of case patients, contact tracing, and testing of people within their networks. Successful implementation of this strategy will require widespread access to testing; substantial efforts are underway to increase SARS-CoV-2 virus testing capacity in the United States and globally. In addition to access (e.g. supply), success of testing strategies will be contingent on the extent to which they are acceptable to patients (e.g. demand). Case identification and contact tracing efforts depend greatly on willingness to test among patients experiencing COVID-19 disease-like symptoms. Those who have tested positive should also be tested again during follow-up care if supplies and laboratory capacity are sufficient, to inform patient isolation strategy.^3^ Patient willingness to seek testing is especially critical because many persons infected with SARS-CoV-2 may experience only mild symptoms: in Italy, 30% of diagnosed cases have been classified as mildly symptomatic,^4^ although such estimates may be an undercount due to the likely lower frequency of test seeking in this group. For other infectious diseases, self-collection procedures have long been practiced,^5,6^ been identified as highly acceptable and preferred to in-clinic procedures,^7^ and having diagnostic metrics comparable to healthcare worker specimen collection.^8,9^

Calls for home-based specimen collection or drive-through specimen collection models to address SARS-CoV-2 virus test scale-up have cogently argued that these approaches have the benefit of (1) avoiding burdening hospitals at a critical time, (2) avoiding potential nosocomial infections, (3) likely lowering costs, and (4) potentially achieving rapid scale-up due to laboratory centralization.^10,11^ One additional benefit of home specimen collection might be that supply chain issues, such as stock-outs of swabs or personal protective equipment, could be alleviated if non-traditional specimens such as saliva or non-traditional locations such as home settings prove sufficient.

Drive-through SARS-CoV-2 virus testing sites already exist,^12^ and a number of laboratories are working to validate home-based self-specimen collection for SARS-CoV-2 testing. Protocols for the self-collection of specimens at home for SARS-CoV-2 testing are currently being explored.^13^ These protocols involve persons being mailed specimen collection materials and instructions, self-collection of specimens at home, and return of specimens to a central laboratory using a supplied mailer. We conducted an online survey to assess patient willingness to use the following SARS-CoV-2 testing modalities for clinical care: home-based specimen collection, drive-through testing, and clinic-based testing. We hypothesized that persons would be more willing to use home-based and drive-through specimen collection modalities compared to clinic-based modalities.

## METHODS

We recruited potential participants using online social media advertisements from March 27th to April 1st, 2020. To be eligible, respondents had to be 18 years of age or older. Given the disproportionate impact of COVID-19 on communities of color, on the final day of data collection eligibility criteria were adjusted to screen out non-Hispanic White respondents in an effort to increase minority representation in the sample. Participants completed an online survey after being recruited from social media sites with banner advertisements requesting participation in COVID-19 survey research. Survey measures included previously published demographic items,^14^ COVID-19 disease knowledge,^15^ COVID-19 disease stigma items,^15^ and a list of COVID-19 disease symptoms based on several sources.^16-18^ We developed a series of questions about willingness to use different testing modalities, each rated with a five-point Likert scale (1-Strongly disagree to 5-Strongly agree). The questions were based on home test willingness questions we have previously used in HIV prevention research.^19^ Definitions for each testing modality were: “A home saliva sample would involve you spitting in a tube and sending it to a certified laboratory,” “A home throat swab would involve you using a throat swab and sending it into a certified laboratory,” “A drive-through site for throat swab would involve your traveling to a drive-through facility in your car to have a healthcare worker collect the swab,” and “A laboratory throat swab would involve your traveling to a laboratory facility in a clinic or private laboratory to have a healthcare worker collect the swab.” Other questions assessed whether persons rated themselves as more likely to seek testing if the option to collect specimens at home for mail-in testing were available.^18^ The full text of survey items can be seen in Supplement 1. All participants completed a written electronic consent procedure, and study procedures were approved by the Emory University IRB.

Figure 1 displays a flowchart of study participation. Figure 2 displays proportion of respondents selecting each willingness option. Table 1 displays demographics and related variables by mean Likert scale values, using Kruskall-Wallace tests for non-ordered categorical variables and Spearman-rank tests for ordinal variables, adjusted for multiple testing using Holm-Bonferroni corrections. Cohen’s *d* effect size was used to guide interpretation of effect size, using standard interpretations. Study reporting follows STROBE criteria for cross-sectional studies.^20^

**Figure 1.**
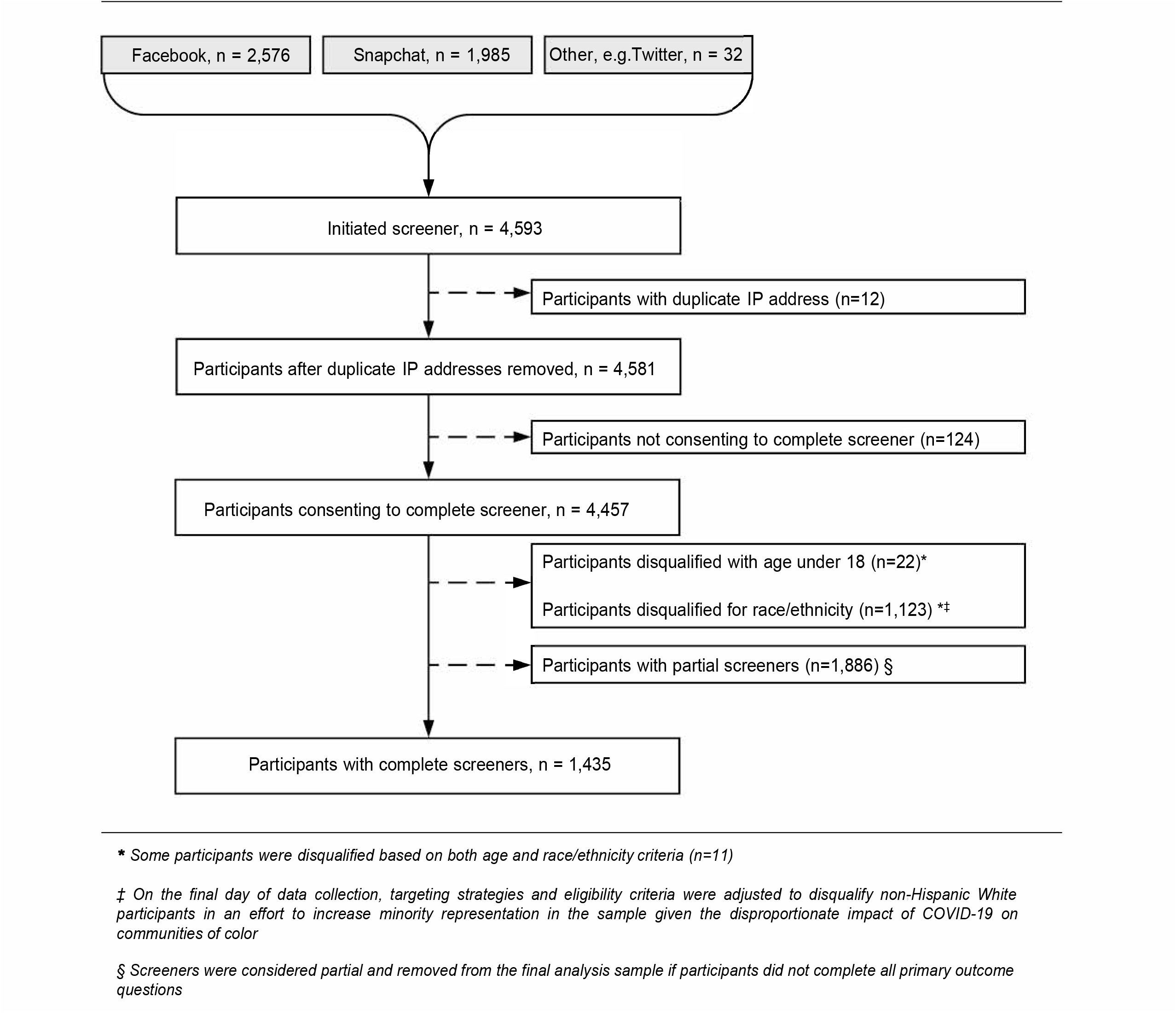
Study participation flowchart.

**Figure 2.**
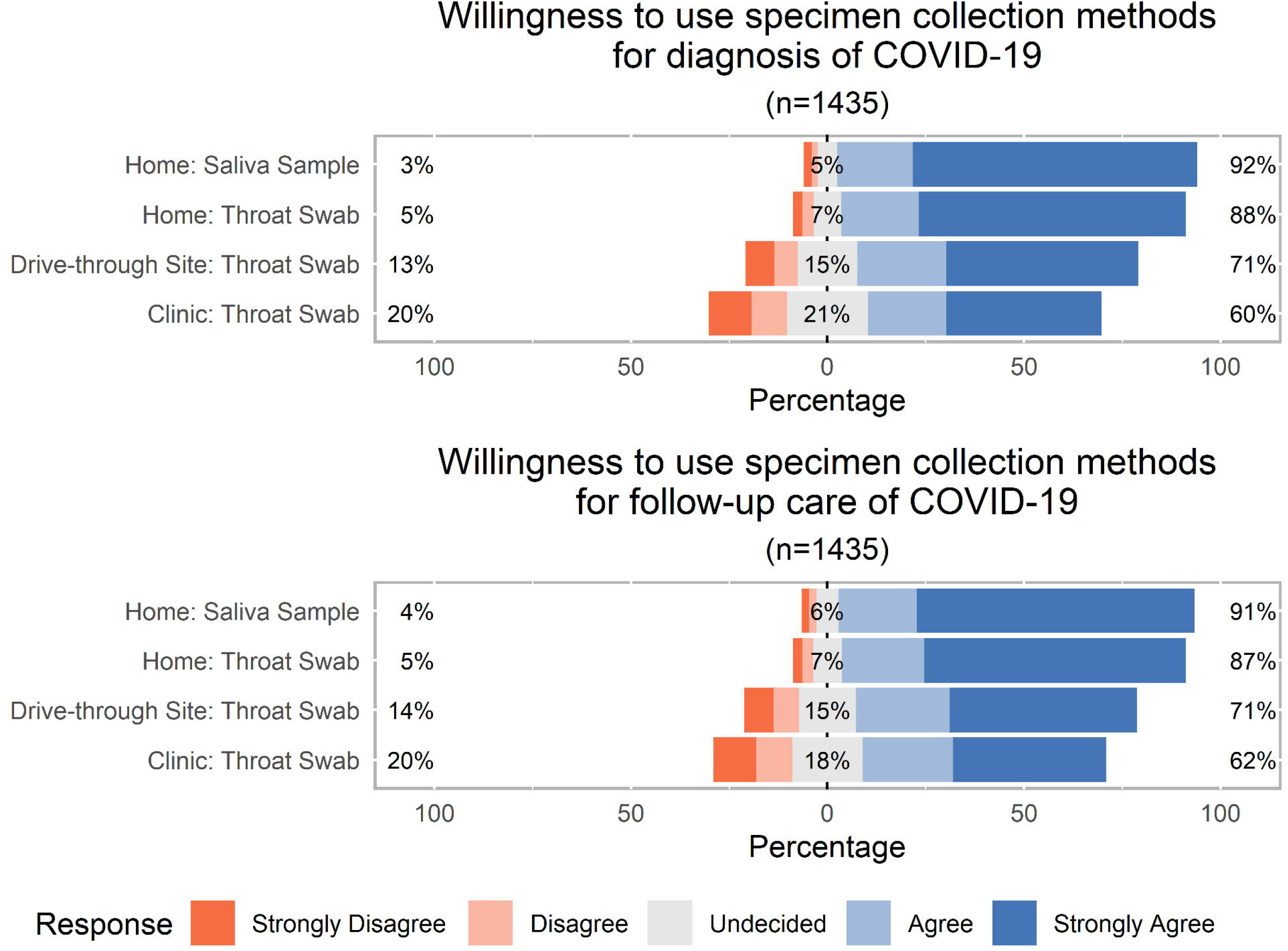
Willingness to seek laboratory testing for SARS-CoV-2 under different specimen collection scenarios * For home specimens, instructions clarified that specimens would be collected at-home and mailed to a central laboratory for testing

**Table 1.**
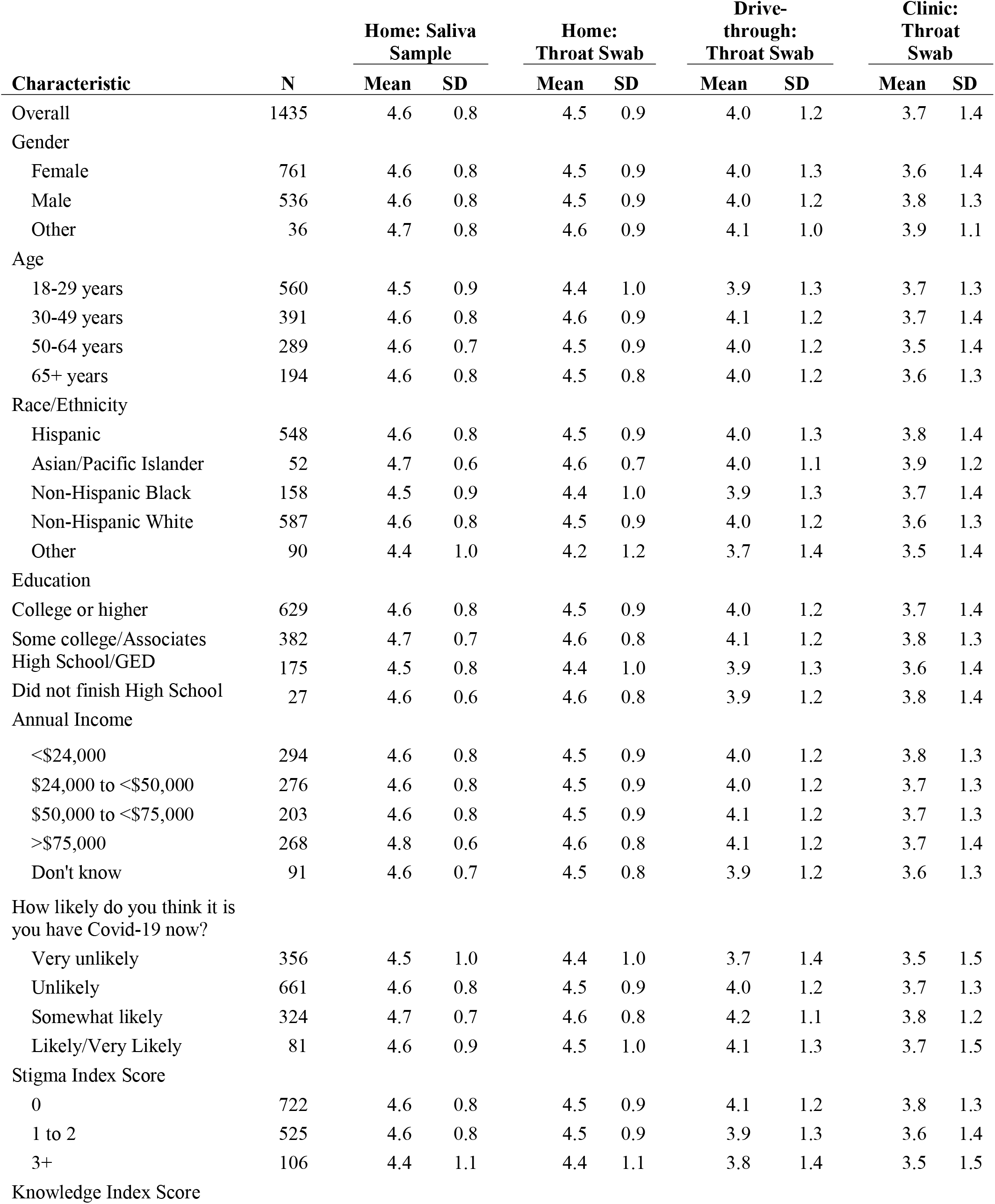
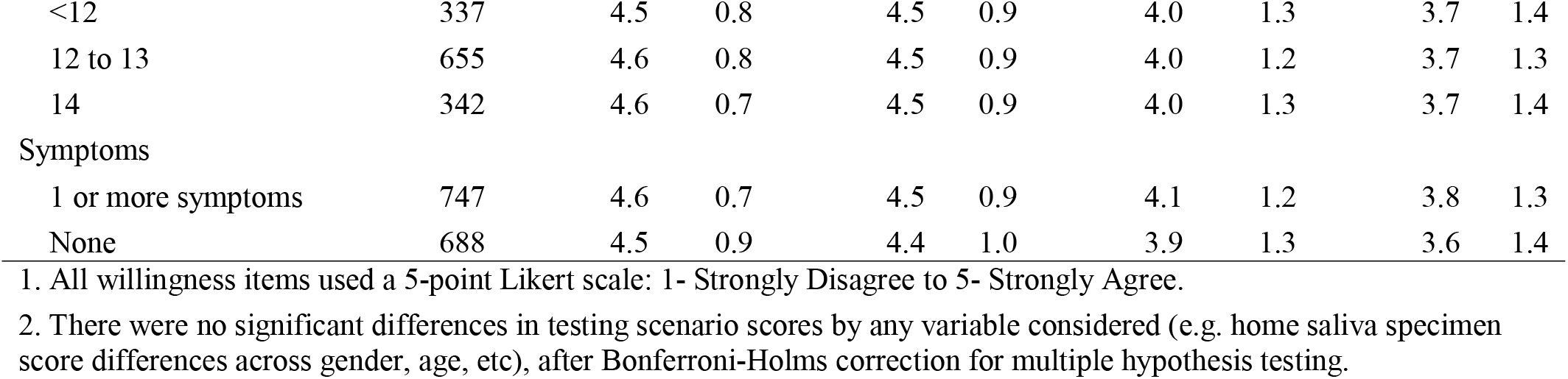
Willingness to seek testing for SARS-CoV-2 if feeling ill, by sociodemographic and COVID-19 factors

## RESULTS

From 4,593 persons initiating the survey screener, 1,260 were ineligible, 1,886 did not consent or provided only partial survey responses, and 1,435 completed all willingness items for the analysis dataset (Figure 1). The sample was 39% (n=560) aged 18-29, 27% (391) aged 30-49, 20% (289) aged 50-64, and 14% (194) aged 65 or older. Females comprised 57% (761), males 40% (536), and other gender identity 3% (36). Overall 41% (587) were non-Hispanic White, 38% (548) were Hispanic, 11% (158) were non-Hispanic Black, 4% (52) were Asian/Pacific Islander, and 6% (90) were Native American/Alaska Native or identified as mixed race or other non-Hispanic. COVID-19 knowledge was high with 75% (997) answering at least 12 of 14 knowledge questions correctly, and COVID-19 stigma was moderate with 46% (631) answering at least one of four stigma questions in a stigmatizing direction. A majority of 72% (1,017) thought they were unlikely to have COVID-19, although 52% (747) reported 1 or more of a broad range of potential COVID-19 symptoms.

Home specimen collection solutions were most preferred with 92% (1314/1435) of participants agreeing or strongly agreeing that they would provide a saliva specimen, and 88% (1258/1435) agreeing that they would provide a throat swab (Figure 2). There was attenuated willingness for drive through swab testing (71%, 1026/1435), and substantially attenuated willingness for clinic or laboratory throat swab (60%, 854/1435). Differences in mean willingness scores across testing modalities were all significant (p<.001), with very small effect size for home saliva testing compared to home throat swab testing (*d*=0.12), medium effect size for home saliva testing compared to drive-through testing (*d*=0.55), and large effect size for home saliva testing compared to clinic-based testing (*d*=0.81). We found highly similar willingness to seek testing for COVID-19 follow-up care (Figure 2), and identical significance and effect size findings (Supplement 2). Willingness to seek testing for diagnosis and care within each testing modality was remarkably consistent across all covariates in the analysis, with no differences across age groups, race/ethnicities, COVID-19 stigma scores, COVID-19 knowledge scores, or COVID-19 symptomology (Table 1).

To directly assess potential behavioral change associated with different home care testing modalities, we asked participants whether they would be more likely, no different, or less likely to seek testing for COVID-19 disease if at-home specimen collection options were available. Relative to availability of a drive-through modality, 65% (933) noted they would be more likely to test if at-home specimen collection were available, 32% (459) noted no difference, and 3% (43) noted lower likelihood. Relative to availability of a clinic- or lab-based modality, 68% (970) noted they would be more likely to test if at-home specimen collection were available, 29% (418) noted no difference, and 3% (47) noted lower likelihood (results not reported in table).

## DISCUSSION

Across a diverse sample of 1,435 participants, one-third more persons reported that they would be willing to collect specimens at home for SARS-CoV-2 testing if they experienced illness, compared to clinic-based testing. There was a hierarchy of willingness to test for SARS-CoV-2 that was decreased as the required degree of contact with healthcare systems increased: home testing was most preferred, followed by drive-through testing, and then by laboratory or clinic-based testing. If reported willingness approximates actual willingness, a difference of this magnitude has considerable public health and clinical care implications. One indicator that the hypothetical may approach actual behavior is that participant preferences were consistent across COVID-19 symptomology levels: persons currently experiencing COVID-19 related symptoms reported similarly lower willingness to seek drive-through and clinic-based SARS-CoV-2 testing as persons not currently experiencing symptoms. Preference differences were also constant across a wide variety of sociodemographic variables, which is important to note, considering the differential impact of SARS-CoV-2 on elderly persons^21^ and on African-Americans, as reported in media and confirmed by coroner’s offices in Louisiana, Chicago, and Michigan.^22^

There are currently vast differences in how countries and jurisdictions are handling testing due to supply limitations. In Iceland, testing has been widely provided as a strategy to combat epidemic spread, and not surprisingly this appears to be substantially contributing to their control of epidemic spread.^23^ At-home self-collection of specimens is one of several options worthy of exploration to achieve similar gains in other settings. Home-based and drive-through testing strategies are promising in part because they may allow for rapid scale-up of newly validated approaches that may relieve supply chain problems. For instance, one laboratory is working to validate testing with saliva specimens, which would relieve current stockouts of nasopharyngeal swab supplies.^13^ It is clear that, if sufficient laboratory capacity and supplies are available, increased testing using at-home specimen collection is critical for public health response for three reasons. First, it would facilitate increased initiation of contact tracing, a tool known to limit epidemic spread, by identifying people with mild symptoms and allowing public health authorities to test close contacts. Second, it would reduce nosocomial infections. Third, it would facilitate improved self-management, because mild and moderate COVID-19 symptoms are nonspecific. Persons receiving a formal SARS-CoV-2 diagnosis are likely to perform self-isolation activities with substantially more rigor than persons whose actions are informed only by their mild symptoms. Conversely, those determined to be uninfected would be anticipated to have reduced anxiety, and be able to continue with their lives without an unnecessary isolation period.

Given our finding that people were more willing to test with home specimen strategies, making such an option available might allow for earlier informed discussions with a clinician via an office visit or telemedicine regarding the optimal next steps in their care. This is especially relevant given media reports, confirmed by local health authorities, of the substantial increases of persons found dead in their homes in some cities in the United States compared to historic averages. In Detroit, there were more than 150 persons founds dead in their homes first 10 days of April compared to around 40 during that same period in the three years prior.^24^ In New York City in early April 2020, a spokesperson for the Department of Health confirmed that around 200 deaths per day have been observed in homes, compared to 20-25 deaths per day in 2019.^25^ It is likely that many of the deceased did not have an opportunity to receive clinical care, a problem that could potentially be mitigated through more wide-spread and easily accessible testing.

Other authors have previously called for SARS-CoV-2 home testing, but mainly for its social distancing and reduced healthcare system burdens.^12,26^ Such calls can and have equally supported drive-through facilities.^12^ But our findings indicate home collection was even substantially preferred to drive-through methods, with over 20% more persons indicating willingness to complete a home test compared to a drive-through test. Drive-through testing venues may achieve benefits of viral transmission control, but have lower benefits for increasing the demand for testing. These results are aligned with previous work that has found home specimen collection a highly preferred method of seeking clinical care, ^19,27,28^ and can be understood as part of an already ongoing move towards remote care facilitated by at-home specimen collection. ^29-32^

This national online survey study has a number of limitations. Participants volunteered to take an online survey regarding COVID-19, potentially skewing willingness values higher than among the general population. We do not think, however, that this would produce bias in the relative levels of support for the testing options presented. The convenience sample may not represent the broader population in other ways, although the consistent and strong differences in preference across categories and item types indicates this would likely have little influence on study results.

We found strong preferences for home testing options. Providing a home testing option is consistent with social distancing strategies and also patient-centered care strategies demonstrated to improve patient adherence to clinician-recommendations. Home specimen collection and central laboratory testing can ease supply chain problems, and be quickly scaled up for contact tracing use by public health authorities. Such home testing methods should be validated as soon as possible, and brought to scale by clinicians and health systems.

## Data Availability

Deidentified data and explanatory documentation will be shared based on written request, and after review and approval of study investigators. Access will be granted after approval by the lead investigators of an analysis proposal and execution of a signed data sharing agreement.

## CRediT authorship contribution statement

Aaron J. Siegler: Conceptualization, Methodology, Writing - original draft, Writing - review & editing. Eric Hall: Formal analysis, Writing - review & editing. Nicole Luisi: Data curation, Writing - review & editing. Maria Zlotorzynska: Project administration, Writing - review & editing. Gretchen Wilde: Writing - review & editing. Travis Sanchez: Writing - review & editing. Heather Bradley: Writing - review & editing. Patrick S. Sullivan: Conceptualization, Writing - review & editing.

## Declaration of interests

All authors have no conflicts of interest to declare.

## Acknowledgements

We appreciate and acknowledge the contributions of our study participants. All authors had full access to study data, and AJS had final responsibility for the decision to submit for publication. The study was facilitated by the Center for AIDS Research at Emory University ( P30AI050409). The content is solely the responsibility of the authors and does not necessarily represent the official views of the National Institutes of Health.

## Notes

### Competing Interest Statement

The authors have declared no competing interest.

